# Correlation between leukocyte phenotypes and prognosis of amyotrophic lateral sclerosis: a longitudinal cohort study

**DOI:** 10.1101/2021.10.05.21264570

**Authors:** Can Cui, Caroline Ingre, Li Yin, Xia Li, John Andersson, Christina Seitz, Nicolas Ruffin, Yudi Pawitan, Fredrik Piehl, Fang Fang

## Abstract

**Background:** Immune response changes have been reported in amyotrophic lateral sclerosis (ALS), but their clinical relevance remains undetermined. Therefore, we aim to evaluate the relationships between blood leukocyte subpopulations and prognosis of ALS.

**Methods:** A longitudinal cohort of 288 ALS patients with up to 5 years of follow-up during 2015-2020 were recruited at the only tertiary referral center for ALS in Stockholm, Sweden. Routine differential leukocyte counts, and determination of lymphocyte subpopulations including an extended T cell panel with flow cytometry, collected at diagnosis and at regular intervals thereafter. The primary outcome was risk of death (alternatively use of invasive ventilation) after diagnosis of ALS. The secondary outcomes included repeatedly measured functional status - through Amyotrophic Lateral Sclerosis Functional Rating Scale-revised (ALSFRS-R) score and disease progression rate. Cox model was used to evaluate the associations between leukocytes and risk of death. Generalized estimating equation model (GEE) was used to assess the correlation between leukocytes and ALSFRS-R score and disease progression rate.

**Results:** The counts of leukocytes, neutrophils and monocytes increased gradually over time since diagnosis and were negatively correlated with ALSFRS-R score, but not associated with risk of death or disease progression rate. Focusing on lymphocyte subpopulations, increasing counts of natural killer (NK) cells (HR=0.61, 95% CI= [0.42-0.88] per SD increase) and proportions of Th2-diffrentiated CD4+ central memory (CM) T cells (HR=0.64, 95% CI= [0.48-0.85] per SD increase) were correlated with a lower risk of death. Increasing proportions of CD4+ effector memory cells re-expressing CD45RA (EMRA) T cells (HR=1.39, 95% CI= [1.01-1.92] per SD increase) and CD8+ T cells (HR=1.38, 95% CI= [1.03-1.86] per SD increase) were associated with a higher risk of death. None of the lymphocyte subpopulations was correlated with ALSFRS-R score or disease progression rate.

**Conclusion:** Our findings suggest a dual role of immune responses in ALS prognosis, where neutrophils and monocytes primarily reflect functional status whereas NK cells and different T lymphocyte populations act as prognostic markers for survival. The findings also provide insights for cell-specific treatment for ALS.

**Funding:** This work was supported by the European Research Council (ERC) Starting Grant (MegaALS, No. 802091), the Swedish Research Council (No. 2019-01088), Karolinska Institutet (Strategic Research Area in Epidemiology and Senior Researcher Award), and China Scholarship Council.

## Introduction

Amyotrophic lateral sclerosis (ALS) is a rare but devastating neurodegenerative disease. Although a genetic cause has been demonstrated for some cases of ALS, the etiology remains unknown for most of the patients with ALS. There is currently no cure or effective treatment available for ALS. A range of potential disease mechanisms have however been proposed, with potential for informing on novel therapeutic targets^1^.

Neuroinflammatory features, including local glial activation and T cell infiltration in the central nervous system (CNS), are well documented in ALS^2^. Animal studies have demonstrated that altering the function of microglia and infiltrating T cells to the CNS affects disease progression in experimental ALS^3-5^. Human studies have also documented signs of systemic immune activation in ALS patients, compared with healthy controls, suggesting that peripheral immune activation may play a role in human ALS through the peripheral-central neuroimmune crosstalk^6,7^. Most of the previous studies^7-9^, however, are not population-based and have limited sample size, raising the concern of potentially insufficient internal (e.g., selection bias and chance finding) and external (e.g., lack of generalizability) validity. Further, few studies have recorded immune cells longitudinally after ALS diagnosis, as most previous studies relied on a single cross-sectional measurement^10,11^.

The main purpose of this study was to determine cellular immune changes occurring over time since diagnosis of ALS in a large community-based sample. For this purpose, we enrolled a longitudinal cohort of ALS patients, representing a large proportion of the entire ALS population in Stockholm, Sweden, to 1) describe the temporal changes of peripheral leukocytes over time since ALS diagnosis, 2) assess the association of immune cell dynamics with the risk of death after ALS diagnosis, and 3) evaluate the correlations of different cell populations with the functional status and disease progression rate of ALS.

## Methods

### Study cohort

The Swedish Motor Neuron Disease (MND) Quality Registry was established in 2015, collecting information on clinical characteristics, biological test results, and quality of life outcomes from >80% of MND patients in Sweden^12^. Since 2017, the MND Quality Registry has included 99% of MND patients in the Stockholm area among whom 97.1% are diagnosed with ALS. All ALS diagnoses were made by a specialist in neurology and followed up by a neuromuscular specialist, and met the diagnostic requirement of definite, probable, probable laboratory-supported, or possible ALS according to the revised El Escorial criteria^13,14^. To ensure the accuracy of diagnosis, all patients in the registry are re-evaluated annually to update diagnosis, whenever needed.

Through the MND Quality Registry, we first identified 420 patients with ALS diagnosed from the start of the registry until October 7^th^, 2020, in Stockholm. We reviewed the medical records of these patients to identify information on peripheral leukocyte populations (i.e., differential leukocyte counts). During this process, we excluded 12 patients who were not diagnosed at the Karolinska University Hospital – the only tertiary referral center for ALS in Stockholm, three patients with unknown time of symptom onset, 82 patients lacking leukocyte counts, and 35 patients with counts outside of the stipulated observation period (i.e., from three months before date of diagnosis until October 7^th^, 2020). The final analysis cohort included 288 patients (68.6%), with at least one recorded differential leukocyte counts during the observation period. The included patients did not differ significantly in terms of demographic and clinical characteristics from the excluded patients (**Table 1**). About half of the patients had a single measurement of leukocytes whereas the other half had been sampled two or more times.

**Table 1.** Characteristics of the 288 patients with amyotrophic lateral sclerosis (ALS) included in the study, compared with the entire population of ALS patients during the study period in Stockholm, Sweden

| Characteristics | Patients included in the study (N=288) | All patients in Stockholm (N=420) | P value for difference* |
| --- | --- | --- | --- |
| **Sex, N (%)** |  |  | P=0.48 |
| Female | 134 (47%) | 201 (48%) |  |
| Male | 154 (53%) | 219 (52%) |  |
| **Age at diagnosis, years** |  |  | P=0.02 |
| Median (Q1, Q3) | 65 (56, 71) | 66 (57, 72) |  |
| **Diagnostic delay, months** |  |  | P=0.94 |
| Median (Q1, Q3) | 12.30 (7.88, 19.93) | 12.35 (7.59, 20.54) |  |
| **Gene mutation, N (%)<sup>+</sup>** |  |  | P=1.00 |
| *SOD1* | 7 (2.88%) | 9 (2.56%) |  |
| *C9orf72* | 22 (9.05%) | 30 (8.55%) |  |
| Other | 4 (1.65%) | 5 (1.42%) |  |
| **Site of onset, N (%)** |  |  | P=0.26 |
| Limb | 182 (63%) | 250 (60%) |  |
| Bulbar | 78 (27%) | 118 (28%) |  |
| Other | 20 (7%) | 32 (8%) |  |
| Missing | 8 (3%) | 20 (5%) |  |
| **Family history, N (%)** |  |  | P=0.19 |
| Yes | 19 (7%) | 30 (7%) |  |
| No | 144 (50%) | 201 (48%) |  |
| Not clear | 3 (1%) | 7 (2%) |  |
| Missing | 122 (42%) | 182 (43%) |  |

| N. of measurements for cell count (%) |  |  | - |
| --- | --- | --- | --- |
| One | 146 (51%) | - |  |
| Two | 75 (26%) | - |  |
| Three | 35 (12%) | - |  |
| Four or more | 32 (11%) | - |  |

\*P value for the differences between patients included in the study and patients not included in the study; Wilcoxon rank sum test was used for the comparison of continuous variables whereas Chi-square test was used for the comparison of categorical variables.
†Results available for 243 of the 288 patients included in the study, and 351 of the entire 420 patients in Stockholm.

Among the ALS patients diagnosed at the Karolinska University Hospital and with a date of symptom onset (N=405), we further performed flow cytometry in 92 patients (“FlowC cohort”) to determine lymphocyte subpopulations (i.e., T, B and natural killer [NK] cells) as well as an extended T lymphocyte panel “FITMaN” - an internationally standardized panel reported by the Flow Immunophenotyping Technical Meeting at NIH^15^. Compared to the main study cohort, patients of the FlowC cohort were slightly younger and more likely to have a limb onset (**eTable 1**). In both the main and FlowC cohorts, we followed the ALS patients from date of diagnosis or first cell measurement (differential leukocyte counts or FlowC), whichever came later, until occurrence of the outcome of interest (i.e., death or use of invasive ventilation) or October 7^th^, 2020, whichever came first.

### Blood samples and flow cytometric analysis

All sample processing and analyzing procedures were according to the validated protocol at the Departments of Clinical Chemistry (differential leukocyte counts) and Clinical Immunology and Transfusion Medicine (FlowC), Karolinska University Hospital. All analyses were performed during daytime, within 24 hours of sampling. Differential leukocyte counts were done on a Sysmex XN-9000 (Sysmex, Kobe, Japan). FlowC was implemented in clinical routine based on the standardized phenotyping panel by the Human Immunophenotyping Consortium with a set of defined 8-color antibody cocktails^15^. The experiments were performed on a triple-laser Beckman Coulter Gallios and analyzed by Kaluza Software (Beckman Coulter, Brea, CA).

Differential leukocyte counts included neutrophils, lymphocytes, monocytes, eosinophils, and basophils. We did not include eosinophils and basophils in the analysis as they in most cases were low to undetectable. The FITMaN panel included measures of 23 lymphocyte subpopulations, including 1) counts of B cells, NK cells, and T cells; 2) %s of CD4^+^ and CD8^+^ T cell subtypes (i.e., naive, central memory [CM], effector memory [EM], and effector memory cells re-expressing CD45RA [EMRA] T cells based on CCR7 and CD45RA expression, as well as Th1, Th2, and Th17 of CM and EM CD4^+^ T cells based on CXCR3 and CCR6 expression); and 3) subtypes of activated CD4^+^ and CD8^+^ T cells based on the expression of HLA-DR and CD38 (i.e., CD4^+^HLA-DR^+^CD38^-^ cells, CD4^+^HLA-DR^+^CD38^+^ cells, CD8^+^HLA-DR^+^CD38^-^ cells, and CD8^+^HLA-DR^+^CD38^+^ cells). T cells were gated from a lymphocyte (FCS/SSC) gate as cells expressing CD45 and CD3. The unit of cell count was 10^9/L whereas the %s were expressed as the proportions of the lower-level immune cell populations out of the upper-level immune cells. The reference values for FlowC were reported in the form of 5^th^ to 95^th^ percentiles using reference normal ranges obtained from 50 healthy adults. All cell counts and %s were retrieved from patients’ clinical files.

### Outcomes of interest

The primary study outcome was risk of death or use of invasive ventilation after ALS diagnosis, identified from the MND Quality Registry. The secondary study outcomes included functional status measured through the Amyotrophic Lateral Sclerosis Functional Rating Scale – revised (ALSFRS-R) and disease progression rate. ALSFRS-R is a questionnaire-based scale that measures the motor function and disease severity of ALS patients and is considered the gold standard measure of disability progression^16^. Higher ALSFRS-R score indicates better functional status. We acquired information on all available ALSFRS-R scores for the ALS patients from the MND Quality Registry. Progression rate measures the rate of ALSFRS-R decline and was calculated by dividing the difference between 48 (the full score) and measured ALSFRS-R score at a specific time point by the time difference between time of symptom onset to the measurement time of ALSFRS-R (in months). Progression rate is an independent prognostic predictor for ALS^17^.

### Statistical analysis

We first calculated the mean levels of measured cell populations among ALS patients, by summarizing all measurements from three months before diagnosis until end of follow-up. To visualize the temporal patterns of the cell populations, we drew a trajectory line of all measurements for each cell type and each patient. We then used the locally estimated scatterplot smoothing (LOESS) curves with 95% confidence intervals (CIs) to show the temporal pattern of the predicted median level of each cell type after ALS diagnosis. If the CIs did not overlap with the normal ranges of the cell populations, we considered the observed levels among ALS patients to be statistically deviant from normal ranges. We also used linear mixed model to assess the within-individual temporal changes of cell populations after ALS diagnosis. In this analysis, we included a random intercept to account for the initial differences between individuals and adjusted for age at diagnosis and sex. We first analyzed all ALS patients together, then separated by site of onset and presence of *C9orf72* expansions. Patients with *C9orf72* expansions have been suggested to demonstrate a different immune phenotype compared with ALS patients without such expansions^18,19^.

We next used Cox model to derive hazard ratio (HR) and 95% CI to assess the association of different cell populations with the risk of death, after adjustment for other prognostic indicators of ALS including age at diagnosis, sex, site of onset, diagnostic delay, ALSFRS-R score, time difference between the measure of ALSFRS-R score and diagnosis, body mass index (BMI), and time difference between the measure of BMI and diagnosis. We obtained information on BMI from the MND Quality Registry. We used time since diagnosis as the underlying time scale and cluster robust variance estimation to account for dependence of repeated measurements. In this analysis, we log-transformed the numbers of leukocytes, neutrophils, lymphocytes and monocytes, as well as %s of CD4^+^ EM cells, CD4^+^ EMRA cells, Th2 of CD4^+^ EM cells, Th17 of CD4^+^ EM cells, Th2 of CD4^+^ CM cells, CD8^+^ T cells, CD8^+^ CM cells, CD4^+^HLA-DR^+^CD38^-^ cells, CD4^+^HLA-DR^+^CD38^+^ cells, CD8^+^HLA-DR^+^CD38^-^ cells, and CD8^+^HLA-DR^+^CD38^+^ cells, to achieve a better normal distribution. Values of other cell populations were used as is (i.e., without transformation). For all markers, we estimated the effect size per standard deviation (SD) increase. Because some patients had their first cell measurements after diagnosis, we also took into account left truncation in all analyses.

The proportional hazards assumption of the Cox model was assessed using the Schoenfeld residual test. After stratifying the analysis by site of onset which deviated from the assumption, the assumption became satisfied for all other variables. As HRs obtained in the stratified analysis were nearly the same as those obtained without stratification, for simplicity and consistency, we reported all findings from the original models (without stratification by site of onset).

Among the 288 patients, 57 (19.8%) were diagnosed before the start of the MND Quality Registry. In a sensitivity analysis, we excluded these patients with the aim to see if the results obtained in the main analysis would pertain to a cohort of incident patients. This analysis was not performed for the FlowC cohort as all the 92 patients were incident patients. We also conducted a sensitivity analysis by only focusing on the first cell measure of each patient to examine whether the main results would differ after removal of repeated measurements. We then performed another sensitivity analysis by excluding patients with *C9orf72* expansions.

We further used a generalized estimating equation (GEE) model to assess the correlations of cell populations with functional status (i.e., ALSFRS-R score) and disease progression rate measured at the same time as the cell markers. The GEE model considers the correlations between repeated measurements within the same individual. Like the survival analysis above, we used log-transformed values for some cell measures whereas original values for others and estimated the effect size per SD increase of the cell measures, after adjustment for age at diagnosis and sex.

Finally, to assess whether the results on cell measures and the study outcomes would be influenced by ongoing infections, we performed another sensitivity analysis after excluding all measurements taken with the presence of an ongoing infection. This was done by reviewing clinical records of all patients with a high count of leukocyte (>8.8×10^9^/L) for diagnosis of any infectious disease, tests for infection, and patient-reported infectious symptoms.

All analyses were performed using R 3.6.2. A two-sided p value of <0.05 was considered statistically significant. To correct for multiple testing, we also computed the Benjamini-Hochberg false discovery rate (FDR)^20^.

## Results

### Leukocytes and lymphocyte subtypes in ALS

**eTable 2** shows the distribution of leukocyte populations (N=288 patients) and lymphocyte subpopulations (N=92 patients) across all measures after ALS diagnosis. The vast majority of the cell populations were within the normal range, except for CD8^+^ CM cells, CD4^+^HLA-DR^+^CD38^+^ cells and CD8^+^HLA-DR^+^CD38^+^ cells, which were above the normal range.

In the main cohort, the levels of leukocytes, neutrophils and monocytes increased progressively over time, especially from 20 months after diagnosis onward (**Figure 1**). These trends were statistically significant, with or without adjustment for age and sex, and remained statistically significant after correction for multiple testing (**Table 2**). In contrast, no clear temporal trend was noted for lymphocytes. These results remained largely similar when stratifying the patients by site of onset or presence of *C9orf72* expansions (**eFigure 1**).

**Figure 1.**
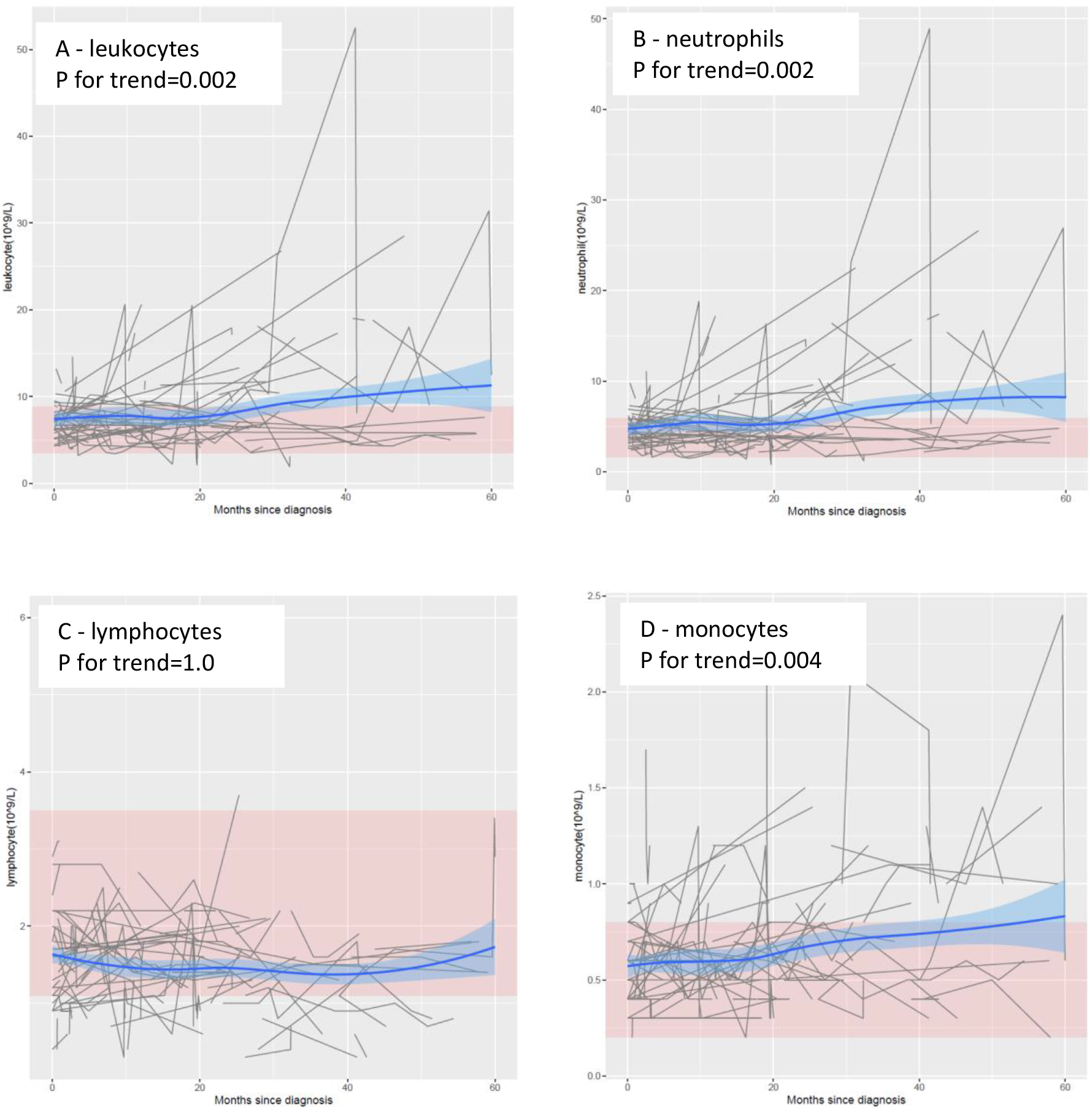
Mean levels of leukocyte populations after a diagnosis of amyotrophic lateral sclerosis (ALS). The black lines show measured levels of leukocyte populations for each patient. The blue lines and shadow areas show the mean levels of leukocyte populations with 95% confidence intervals. Pink areas indicate normal range. P for trend shows the P value of within-individual temporal change of each cell population after taking into account the relatedness of repeated measurements.

**Table 2.** Temporal changes of leukocyte populations after diagnosis of amyotrophic lateral sclerosis (ALS), a cohort study of 288 patients with ALS in Stockholm, Sweden*

| Cell type | Unadjusted |  |  | Adjusted<sup>+</sup> |  |  |
| --- | --- | --- | --- | --- | --- | --- |
|  | Coefficient | P value | FDR | Coefficient | P value | FDR |
| Leukocyte (10<sup>9</sup>/L) | 0.19 | **0.01** | **0.01** | 0.22 | **2.4E-03** | **4.7E-03** |
| Neutrophil (10<sup>9</sup>/L) | 0.18 | **3.6E-03** | **0.01** | 0.21 | **1.5E-03** | **4.7E-03** |
| Lymphocyte (10<sup>9</sup>/L) | 3.7E-03 | 0.73 | 0.73 | 4.1E-05 | 1.00 | 1.00 |
| Monocyte (10<sup>9</sup>/L) | 0.01 | **0.03** | **0.04** | 0.01 | **4.2E-03** | **0.01** |

\*Linear mixed model was applied to derive the coefficient estimates, per year and p value for trend.
<sup>+</sup>Adjusted for age at diagnosis and sex.
FDR: False discovery rate.

In the FlowC cohort, no clear temporal trend was noted for any lymphocyte subpopulation, although ALS patients demonstrated persistently higher proportions of CD8^+^ CM, CD4^+^HLA-DR^+^CD38^+^ and CD8^+^HLA-DR^+^CD38^+^ cells than the reference ranges (**eFigure 2**). After adjustment for age and sex, there was a decreasing %s of naïve CD4^+^ T cells whereas increasing %s of CD4^+^ EMRA, CD4^+^HLA-DR^+^CD38^-^ and CD8^+^HLA-DR^+^CD38^-^ cells since ALS diagnosis (**eTable 3**). Patients with limb onset had lower levels of CD8^+^ CM and CD4^+^HLA-DR^+^CD38^+^ cells compared with patients with other anatomical disease onset, whereas carriers of *C9orf72* expansions had higher levels of NK cells and T cells, but lower levels of naïve CD4^+^ T cells and CD8^+^ EM cells, than other patients (**eFigure 3**).

### Survival

During a median follow-up of 1.1 years, we observed 163 deaths or use of invasive ventilation among the 288 patients of the main cohort. No association was noted between the level of leukocytes, neutrophils, lymphocytes, or monocytes with risk of death (**eTable 4**). This result did not change after excluding patients diagnosed before the start of the MND Quality Registry, focusing on first cell measure only, or excluding patients with *C9orf72* expansions (**eTable 5**).

In the FlowC cohort, we found higher NK cell counts and %s of Th2-diffrentiated CD4^+^ CM cells to be associated with lower risk of death, whereas higher %s of CD4^+^ EMRA cells and CD8^+^ T cells were associated with higher risk of death (**Figure 2**). These results were largely similar after restricting the analysis to first measure of lymphocytes (data not shown) or after excluding patients with *C9orf72* expansions (**eFigure 4**).

**Figure 2.**
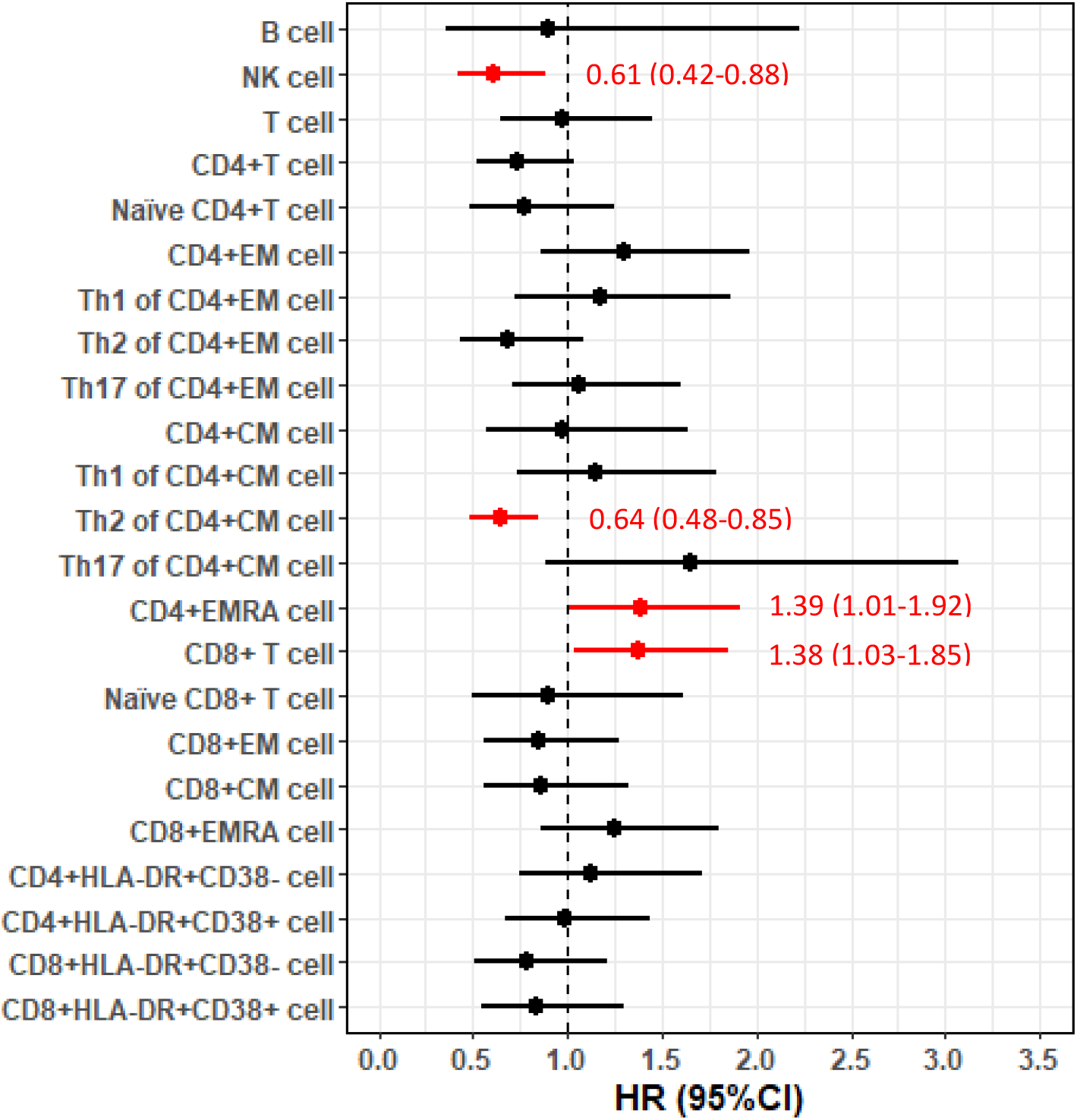
Forest plot of hazard ratios (HRs) and 95% confidence intervals (CIs) for the associations of lymphocyte populations with risk of death after a diagnosis of amyotrophic lateral sclerosis (ALS).

### Functional status and disease progression rate

In the main cohort, a higher level of leukocytes, neutrophils, or monocytes was associated with a lower ALSFRS-R score measured at the time of sampling, whereas no such correlation was evident for lymphocyte counts (**Table 3**). There was, however, no association of leukocytes, neutrophils, lymphocytes, or monocytes with disease progression rate.

**Table 3.** Cross-sectional correlations between leukocyte populations and ALS functional rating scale-revised (ALSFRS-R) score and disease progression rate, a cohort study of 288 ALS patients in Stockholm, Sweden*

| Cell type | ALSFRS-R |  |  | Progression rate |  |  |
| --- | --- | --- | --- | --- | --- | --- |
|  | Coefficient | P value | FDR | Coefficient | P value | FDR |
| Leukocyte (10<sup>9</sup>/L) | -2.80 | **4.0E-03** | **0.01** | 0.02 | 0.74 | 0.74 |
| Neutrophil (10<sup>9</sup>/L) | -3.10 | **1.0E-03** | **4.0E-03** | 0.05 | 0.33 | 0.67 |
| Lymphocyte (10<sup>9</sup>/L) | 1.48 | 0.15 | 0.15 | -0.08 | 0.32 | 0.67 |
| Monocyte (10<sup>9</sup>/L) | -2.75 | **2.0E-03** | **4.0E-03** | -0.03 | 0.52 | 0.69 |

\*Generalized estimating equation model was applied to derive the coefficient estimates and p values, with adjustment for age at diagnosis and sex. ALSFRS-R score ranges from 0 to 48, with higher score showing better motor function status. Progression rate indicates the decline of motor function per month.
FDR: false discovery rate.

In the FlowC cohort, none of the lymphocyte subtypes was associated with ALSFRS-R score or disease progression rate measured at the time of sampling (**eTable 6**).

### Sensitivity analysis for ongoing infection

We excluded 15 patients in the main cohort who had been sampled with the presence of infection. The results on risk of death, ALSFRS-R score and disease progression rate remained similar although some results lost statistical significance (**eTable 7** and **eTable 8**). There was no patient with sampling during ongoing infection in the FlowC cohort.

## Discussion

We here report a longitudinal cohort study of temporal dynamics of white blood cell populations in 288 ALS patients in Stockholm, Sweden. We found higher counts of blood leukocytes, neutrophils and monocytes to be associated with a lower functional status, but not with the risk of death after ALS diagnosis or disease progression rate. In a subsample of 92 patients, we found that higher NK cell counts and proportions of Th2-diffrentiated CD4^+^ T cells were associated with a lower risk of death, whereas higher counts of CD8^+^ T cells and proportions of CD4^+^ EMRA T cells were associated with higher mortality risk.

ALS patients demonstrated gradually increasing counts of leukocytes, neutrophils and monocytes over time since diagnosis. Benjamin et al. also reported increasing counts of leukocytes and neutrophils in ALS patients^21^. Accumulating monocytes have also been shown in the cervical and lumbar spinal cord of ALS model over time^22^. The finding that peripheral leukocytes, neutrophils, and monocytes were correlated to ALSFRS-R, but not progression rate or survival, indicates that these leukocyte subtypes may serve better as markers for functional status than prognosis. A previous study also found CD16 expression on neutrophils and non-classical monocytes to correlate with ALS disease severity^23^. As neutrophils and monocytes have phagocytic function, increased levels of circulating neutrophils and monocytes might indicate enhanced muscle damage, explaining the correlation with deteriorating functional status. It has also been reported that circulating monocytes from ALS patients preferentially differentiate to a M1 proinflammatory phenotype and produce more Interleukin 6 (IL-6) and tumor necrosis factor α (TNF-α), compared with monocytes from healthy controls^24^. Regardless, although it has been proposed that peripheral neutrophils and monocytes are recruited to the CNS through a disrupted brain-blood barrier (BBB) and affect the disease progression of experimental ALS through secreting proinflammatory cytokines and influencing other cells^25-27^, the functional relevance in human ALS is still unclear^22,28^.

We found a higher level of blood-borne NK cells to be associated with a lower risk of death after ALS diagnosis. NK cells are considered a critical part of the innate immunity and function through lysing infected, oncogenic, apoptotic, and MHC class I-deficient cells^29^. Although NK cells are known to penetrate BBB and interact with microglia, astroglia, and neurons^30^, the role of NK cells in disease progression of ALS has rarely been addressed. NK cells may exert a cytotoxic role in the brain by its natural function, as suggested by a study showing that depletion of NK cells prolonged survival in ALS mouse models^31^. As infiltration of NK cells to CNS may lead to a decrease in peripheral NK populations, the protective role of higher peripheral NK cells against risk of death, as observed in the present study, might be partly attributable to a lower level of its infiltration to the CNS. On the other hand, NK cells may indeed also play a neuroprotective role. For instance, findings from experimental autoimmune encephalomyelitis, a model of multiple sclerosis, showed NK cells to suppress neuroinflammation, diminish tissue damage and protect motoneurons^32,33^. Patients with multiple sclerosis have also been shown to experience clinical improvement through NK cell expansions^34^. NK cells have also been shown to exert a protective role in brain by removing viral infection and activated microglia^30^.

We also found that higher Th2 differentiation of CD4^+^ T cells was associated with a lower risk of death after ALS diagnosis, corroborating previous findings of the neuroprotective role of CD4^+^ T cells in ALS^5,35^. Animal studies have suggested Th1 cells and Th17 cells to promote neuroinflammation by producing proinflammatory cytokines and enhancing microglia-mediated neurotoxic effects^36^ whereas Th2 cells and regulatory T cells (Tregs) to suppress neuroinflammation by producing anti-inflammatory cytokines and enhancing microglia-mediated neuroprotective effects^37,38^. Although not statistically significant, our study indeed found a trend for Th1-differentiatated CD4+ CM cells and Th17-differentiated CD4+ CM cells to be positively associated with the risk of death after ALS diagnosis.

A novel finding of the present study is that higher proportions of CD8^+^ T cells and CD4^+^ EMRA T cells were associated with a higher risk of death after ALS diagnosis. The precise underlying mechanisms linking together these cell types with ALS survival are unknown. Previously, CD8^+^ T cells have been found to be present in the spinal cord only at the end stage of ALS^5^. However, such cells have been suggested to have a pathogenic potential in multiple sclerosis and to communicate with mononuclear phagocytes^39,40^. Previous studies also indicated that CD4^+^ EMRA cells could demonstrate cytotoxic features and express the chemokine receptor CX3CR1 in the setting of Dengue virus infection^41^, which are associated with cytotoxic lymphocytes with cytoplasmic granules containing perforin and granzymes^42^. Further studies are clearly needed to study more in detail the functional relevance of CD8^+^ and CD4^+^ EMRA T cells in ALS.

Expansions in *C9orf72*, the most common genetic cause of ALS, have been shown to be associated with immune features^43^, including activation of microglia and elevated levels of peripheral inflammatory cytokines^44^. In our study, although patients with or without *C9orf72* expansions did not differ greatly in terms of major leukocyte populations, patients with *C9orf72* expansions appeared to display changes in certain lymphocyte subpopulations including NK, T, naïve CD4^+^ T and CD8^+^ EM cells compared with patients without such expansions. Because of the relatively small number of patients with *C9orf72* expansions in the study, these results should however be interpreted with caution until validated further.

Our study is the first, to our knowledge, to report the role of a comprehensive list of immune cells, including neutrophils, lymphocytes, monocytes and detailed T cell phenotypes, on the prognosis of ALS. The strengths of the present study are the large number of ALS patients which were representative of all ALS patients in the source population, the rich information on disease characteristics including genetic causes, the long (up to 5 years after diagnosis) and complete follow-up (through the MND Quality Registry), the availability of both routine cell counts and detailed lymphocyte phenotyping, as well as the access to repeated measures over time. The study also has limitations. First, the cohort was heterogeneous in terms of the numbers of cell measurements and the time intervals between measurements. Some concern of indication bias, for instance, due to impact of infections, might exist. We therefore excluded cell measures taken at the time of infection in a sensitivity analysis. The largely similar results obtained in this analysis relieved this concern to some extent. Further, the FITMaN panel does not include B cell subtypes or Treg cells, which await to be studied further. Finally, causal inferences on the functional implications of the reported associations are not possible due to the observational study design.

In conclusion, our findings suggest a dual role of immune responses in ALS prognosis, where neutrophils and monocytes primarily reflect functional status whereas different T lymphocyte populations act as prognostic markers for survival, which also provide insights for cell-based therapy in prolonging survival in ALS.

## Data Availability

The original data are held by the Swedish Motor Neuron Disease Quality Registry and are not publicly available due to Swedish laws. However, any researcher who is interested can access the data by obtaining an ethical approval from regional ethical review board. Detailed information of the Swedish Ethical Review Authority can be found at https://etikprovningsmyndigheten.se. Anonymized data that support the findings of this study are available on reasonable request from the corresponding authors, in agreement with European regulations and Regional Ethical Review Board in Sweden. One source file containing R code for table 2 and 3 has been provided.

## Acknowledgements

This work was supported by the European Research Council (ERC) Starting Grant (MegaALS, No. 802091), the Swedish Research Council (No. 2019-01088), Karolinska Institutet (Strategic Research Area in Epidemiology and Senior Researcher Award), and China Scholarship Council. The funders had no role in the design of the study and collection, analysis, and interpretation of data and in writing the manuscript.

## Online Supplementary Content

**Supplementary Table 1.** Characteristics of the 92 patients with amyotrophic lateral sclerosis (ALS) included in the analysis of FlowC test, compared with the entire population of ALS patients during the study period in Stockholm, Sweden

| Characteristics | Patients included in the analysis (N=92) | All patients in Stockholm (N=420) | P value for difference* |
| --- | --- | --- | --- |
| **Sex, N (%)** |  |  | P=0.29 |
| Female | 49 (53%) | 201(48%) |  |
| Male | 43 (47%) | 219 (52%) |  |
| **Age at diagnosis, years** |  |  | P=0.002 |
| Median (Q1,Q3) | 62 (54, 70) | 66 (57,72) |  |
| **Diagnostic delay, months** |  |  | P=0.59 |
| Median (Q1,Q3) | 11.98 (7.52,19.89) | 12.35 (7.59,20.54) |  |
| **Gene mutation, N (%)** |  |  | P=0.55 |
| *SOD1* | 3 (4.11%) | 9 (2.56%) |  |
| *C9orf72* | 10 (13.70%) | 30 (8.55%) |  |
| Other | 3 (4.11%) | 5 (1.42%) |  |
| **Site of onset, N (%)** |  |  | P=0.03 |
| Limb | 64 (70%) | 250 (60%) |  |
| Bulbar | 20 (22%) | 118 (28%) |  |
| Other | 3 (3%) | 32 (8%) |  |
| Missing | 5 (5%) | 20 (5%) |  |
| **Family history, N (%)** |  |  | P=0.60 |
| Yes | 7 (8%) | 30 (7%) |  |
| No | 35 (38%) | 201 (48%) |  |
| Not clear | 2 (2%) | 7 (2%) |  |
| Missing | 48 (52%) | 182 (43%) |  |
| **N. of measurements for FlowC test (%)** |  |  |  |
| One | 10 (11%) | - |  |
| Two | 23 (25%) | - |  |
| Three | 23 (25%) | - |  |
| Four | 24 (26%) | - |  |
| Five | 10 (11%) | - |  |
| Six | 2 (2%) | - |  |

\*P value for the differences between patients included in the analysis and patients not included in the analysis; Wilcoxon rank sum test was used for the comparison of continuous variables whereas Chi-square test was used for the comparison of categorical variables.
\*Results available for 73 of the 92 patients included in the analysis, and 351 of the entire 420 patients in Stockholm.

**Supplementary Table 2.** Mean levels of leukocyte subpopulations (N=288 patients) and lymphocyte subpopulations (N=92 patients) across all measures

| Cell type | Mean (SD) | Normal range |
| --- | --- | --- |
| Leukocyte (10<sup>9</sup>/L) | 7.8 (3.72) | 3.50-8.80 |
| Neutrophil (10<sup>9</sup>/L) | 5.31 (3.01) | 1.60-5.90 |
| Lymphocyte (10<sup>9</sup>/L) | 1.58 (0.6) | 1.10-3.50 |
| Monocyte (10<sup>9</sup>/L) | 0.58 (0.22) | 0.20-0.80 |
| T cell (10<sup>9</sup>/L) | 1.06 (0.37) | 0.65-1.57 |
| B cell (10<sup>9</sup>/L) | 0.2 (0.17) | 0.08-0.28 |
| NK cell (10<sup>9</sup>/L) | 0.21 (0.1) | 0.10-0.35 |
| CD4+ T cell (%) | 65.32 (11.2) | 44-79 |
| CD4+ naïve T cell (%) | 33.19 (14.54) | 22-62 |
| CD4+ EM (%) | 17.45 (10.76) | 10-47 |
| CD4+ CM (%) | 45.58 (13.02) | 14-48 |
| CD4+ EMRA (%) | 4 (7.39) | 1-9 |
| Th1 of CD4+ EM (%) | 49.19 (13.95) | 22-52 |
| Th2 of CD4+ EM (%) | 15.31 (10.67) | 4-27 |
| Th17 of CD4+ EM (%) | 12.4 (6.58) | 8-28 |
| Th1 of CD4+ CM (%) | 26.53 (5.46) | 13-30 |
| Th2 of CD4+ CM (%) | 26.87 (7.56) | 14-53 |
| Th17 of CD4+ CM (%) | 23.72 (4.91) | 20-37 |
| CD8+ T cell (%) | 30.2 (10.67) | 17-47 |
| CD8+ naïve T cell (%) | 22.27 (14.54) | 10-65 |
| CD8+ EM (%) | 24.64 (11.63) | 4-30 |
| CD8+ CM (%) | **15.81 (9.98)** | 1-12 |
| CD8+ EMRA (%) | 37.37 (18.41) | 18-68 |
| CD4+ HLA-DR+ CD38- (%) | 4.11 (2.85) | 2-11 |
| CD4+ HLA-DR+ CD38+ (%) | **3.24 (1.87)** | ≤2 |
| CD8+ HLA-DR+ CD38- (%) | 6.45 (5.03) | 2-22 |
| CD8+ HLA-DR+ CD38+ (%) | **8.84 (6.61)** | 1-5 |

SD: standard deviation
Bold values indicate deviants from normal range.

**Supplementary Table 3.** Temporal changes of lymphocyte populations after diagnosis of amyotrophic lateral sclerosis (ALS), analysis of 92 patients with FlowC test*

| Cell type | Unadjusted |  |  | Adjusted<sup>+</sup> |  |  |
| --- | --- | --- | --- | --- | --- | --- |
|  | Coefficient | P value | FDR | Coefficient | P value | FDR |
| T cell (10<sup>9</sup>/L) | 0.02 | 0.26 | 0.58 | 0.01 | 0.63 | 0.81 |
| B cell (10<sup>9</sup>/L) | 0.01 | 0.22 | 0.56 | 0.01 | 0.36 | 0.55 |
| NK cell (10<sup>9</sup>/L) | -5.4E-04 | 0.91 | 0.92 | 4.1E-04 | 0.94 | 0.98 |
| CD4+ T cell (%) | 0.89 | 0.03 | 0.19 | 1.07 | 0.01 | 0.06 |
| CD4+ naïve T cell (%) | -0.93 | 0.03 | 0.19 | -1.16 | **0.01** | **0.04** |
| CD4+ EM (%) | -0.06 | 0.88 | 0.92 | 0.01 | 0.99 | 0.99 |
| CD4+ CM (%) | -0.25 | 0.56 | 0.75 | -0.09 | 0.83 | 0.96 |
| CD4+ EMRA (%) | 0.73 | 0.01 | 0.12 | 0.88 | **1.6E-03** | **0.01** |
| Th1 of CD4+ EM (%) | 0.65 | 0.31 | 0.58 | 0.69 | 0.31 | 0.51 |
| Th2 of CD4+ EM (%) | -0.52 | 0.32 | 0.58 | -0.67 | 0.24 | 0.50 |
| Th17 of CD4+ EM (%) | -0.14 | 0.60 | 0.76 | -0.02 | 0.94 | 0.98 |
| Th1 of CD4+ CM (%) | -0.17 | 0.47 | 0.68 | -0.28 | 0.26 | 0.50 |
| Th2 of CD4+ CM (%) | 0.48 | 0.11 | 0.36 | 0.66 | 0.03 | 0.13 |
| Th17 of CD4+ CM (%) | -0.32 | 0.12 | 0.36 | -0.27 | 0.20 | 0.50 |
| CD8+ T cell (%) | -0.37 | 0.33 | 0.58 | -0.50 | 0.20 | 0.50 |
| CD8+ naïve T cell (%) | 0.05 | 0.92 | 0.92 | -0.48 | 0.28 | 0.50 |
| CD8+ EM (%) | -0.64 | 0.13 | 0.36 | -0.29 | 0.51 | 0.73 |
| CD8+ CM (%) | 0.05 | 0.89 | 0.92 | 0.19 | 0.60 | 0.81 |
| CD8+ EMRA (%) | 0.43 | 0.45 | 0.68 | 0.70 | 0.23 | 0.50 |
| CD4+ HLA-DR+ CD38- (%) | 0.21 | 0.07 | 0.31 | 0.35 | **1.7E-03** | **0.01** |
| CD4+ HLA-DR+ CD38+ (%) | -0.06 | 0.42 | 0.68 | 0.02 | 0.84 | 0.96 |
| CD8+ HLA-DR+ CD38- (%) | 0.57 | 0.01 | 0.12 | 0.84 | **2.8E-04** | **0.01** |
| CD8+ HLA-DR+ CD38+ (%) | -0.07 | 0.81 | 0.92 | 0.37 | 0.25 | 0.50 |

\*Linear mixed model was applied to derive the coefficient estimates, per year and p value for trend.
<sup>+</sup>Adjusted for age at diagnosis and sex.
FDR: False discovery rate.

**Supplementary Table 4.** Associations of leukocyte populations with the risk of death after a diagnosis of amyotrophic lateral sclerosis (ALS), a cohort study of 288 patients with ALS in Stockholm, Sweden

| Cell type | HR (95%CI)* | P value | FDR |
| --- | --- | --- | --- |
| Leukocyte (10<sup>9</sup>/L) | 1.16 (0.93-1.45) | 0.18 | 0.36 |
| Neutrophil (10<sup>9</sup>/L) | 1.16 (0.94-1.44) | 0.17 | 0.36 |
| Lymphocyte (10<sup>9</sup>/L) | 0.98 (0.78-1.23) | 0.85 | 0.85 |
| Monocyte (10<sup>9</sup>/L) | 1.07 (0.86-1.32) | 0.55 | 0.73 |

\*Cox model was applied to derive the hazard ratios (HRs) with 95% confidence intervals (CIs) of risk of death, per standard deviation increase of the cell markers, with adjustment for age at diagnosis, sex, site of onset, diagnostic delay, ALSFRS-R score, time difference between the measure of ALSFRS-R score and diagnosis, BMI, and time difference between the measure of BMI and diagnosis.
FDR: false discovery rate.

**Supplementary Table 5.** Sensitivity analyses of the associations of leukocyte populations with risk of death after a diagnosis of amyotrophic lateral sclerosis (ALS)*

| Cell type | HR (95%CI)<sup>1</sup> | HR (95%CI)<sup>2</sup> | HR (95%CI)<sup>3</sup> |
| --- | --- | --- | --- |
| Leukocyte (10<sup>9</sup>/L) | 1.13 (0.90-1.42) | 0.95 (0.76-1.18) | 1.19 (0.95-1.51) |
| Neutrophil (10<sup>9</sup>/L) | 1.15 (0.93-1.41) | 1.02 (0.82-1.26) | 1.17 (0.94-1.46) |
| Lymphocyte (10<sup>9</sup>/L) | 0.90 (0.74-1.09) | 0.94 (0.74-1.18) | 1.02 (0.81-1.29) |
| Monocyte (10<sup>9</sup>/L) | 0.97 (0.79-1.19) | 0.82 (0.65-1.03) | 1.09 (0.88-1.36) |

\*Cox model was applied to derive the hazard ratios (HRs) with 95% confidence intervals (CIs) of risk of death, per standard deviation increase of the cell markers, with adjustment for age at diagnosis, sex, site of onset, diagnostic delay, ALSFRS-R score, time difference between the measure of ALSFRS-R score and diagnosis, BMI, and time difference between the measure of BMI and diagnosis.
<sup>1</sup>Analysis restricted to newly diagnosed ALS patients.
<sup>2</sup>Analysis restricted to the first measurement of cell populations.
<sup>3</sup>Analysis restricted to patients without *C9orf72* mutation.

**Supplementary Table 6.** Cross-sectional correlations between lymphocyte populations and ALS functional rating scale-revised (ALSFRS-R) score and disease progression rate, a cohort study of 92 ALS patients in Stockholm, Sweden*

| Cell type | ALSFRS-R |  |  | Progression rate |  |  |
| --- | --- | --- | --- | --- | --- | --- |
|  | Coefficient | P value | FDR | Coefficient | P value | FDR |
| T cell($10^9/L$) | 0.11 | 0.90 | 0.97 | -0.11 | 0.06 | 0.43 |
| B cell($10^9/L$) | -0.85 | 0.31 | 0.95 | -0.05 | 0.20 | 0.50 |
| NK cell($10^9/L$) | 0.57 | 0.51 | 0.95 | -0.10 | **0.02** | 0.43 |
| CD4+ T cell(%) | -0.06 | 0.96 | 0.97 | -0.04 | 0.32 | 0.62 |
| CD4+ naïve T cell(%) | 0.60 | 0.57 | 0.95 | -0.06 | 0.32 | 0.62 |
| CD4+ EM(%) | 1.17 | 0.22 | 0.95 | 0.06 | 0.22 | 0.51 |
| CD4+ CM(%) | -0.26 | 0.83 | 0.97 | 0.09 | 0.10 | 0.43 |
| CD4+ EMRA(%) | -0.71 | 0.53 | 0.95 | -0.10 | 0.09 | 0.43 |
| Th1 of CD4+ EM(%) | 0.09 | 0.93 | 0.97 | 0.02 | 0.80 | 0.88 |
| Th2 of CD4+ EM(%) | 0.86 | 0.31 | 0.95 | -0.10 | 0.11 | 0.43 |
| Th17 of CD4+ EM(%) | 0.35 | 0.75 | 0.97 | 0.03 | 0.65 | 0.84 |
| Th1 of CD4+ CM(%) | 1.40 | 0.18 | 0.95 | -0.01 | 0.87 | 0.91 |
| Th2 of CD4+ CM(%) | 0.03 | 0.97 | 0.97 | -0.09 | 0.16 | 0.46 |
| Th17 of CD4+ CM(%) | -0.58 | 0.58 | 0.95 | 0.04 | 0.53 | 0.83 |
| CD8+ T cell(%) | 0.40 | 0.73 | 0.97 | 0.07 | 0.15 | 0.46 |
| CD8+ naïve T cell(%) | 0.45 | 0.69 | 0.97 | -0.05 | 0.46 | 0.81 |
| CD8+ EM(%) | -1.06 | 0.37 | 0.95 | 0.02 | 0.79 | 0.88 |
| CD8+ CM(%) | 0.25 | 0.82 | 0.97 | 0.10 | 0.06 | 0.43 |
| CD8+ EMRA(%) | 0.66 | 0.55 | 0.95 | -0.04 | 0.54 | 0.83 |
| CD4+ HLA-DR+ CD38- (%) | -1.75 | 0.07 | 0.95 | -2.6E-03 | 0.97 | 0.97 |
| CD4+ HLA-DR+ CD38+(%) | -1.01 | 0.22 | 0.95 | -0.03 | 0.66 | 0.84 |
| CD8+ HLA-DR+ CD38- (%) | -0.65 | 0.57 | 0.95 | 0.03 | 0.59 | 0.84 |
| CD8+ HLA-DR+ CD38+(%) | -1.43 | 0.10 | 0.95 | -0.02 | 0.78 | 0.88 |

\*Generalized estimating equation model was applied to derive the coefficient estimates and p values, with adjustment for age at diagnosis and sex. ALSFRS-R score ranges from 0 to 48, with higher score showing better motor function status. Progression rate indicates the decline of motor function per month.
FDR: false discovery rate.

**Supplementary Table 7.** Sensitivity analyses of the associations of leukocyte populations with ALS functional rating scale-revised (ALSFRS-R) score and disease progression rate, a cohort study of 273 ALS patients in Stockholm, Sweden*

| Cell type | ALSFRS-R |  |  | Progression rate |  |  |
| --- | --- | --- | --- | --- | --- | --- |
|  | Coefficient | P value | FDR | Coefficient | P value | FDR |
| Leukocyte (10<sup>9</sup>/L) | -1.40 | 0.13 | 0.15 | 0.04 | 0.41 | 0.55 |
| Neutrophil (10<sup>9</sup>/L) | -1.72 | 0.05 | 0.10 | 0.07 | 0.12 | 0.48 |
| Lymphocyte (10<sup>9</sup>/L) | 1.28 | 0.15 | 0.15 | -0.07 | 0.41 | 0.55 |
| Monocyte (10<sup>9</sup>/L) | -1.94 | **4.0E-03** | **0.02** | -0.01 | 0.90 | 0.90 |

\*Generalized estimating equation model was applied to derive the coefficient estimates and p values, with adjustment for age at diagnosis and sex.
ALSFRS-R score ranges from 0 to 48, with higher score showing better motor function status.
Progression rate indicates the decline of motor function per month.
FDR: false discovery rate.

**Supplementary Table 8.** Sensitivity analyses of associations of leukocyte populations with the risk of death after a diagnosis of amyotrophic lateral sclerosis (ALS), a cohort study of 273 ALS patients in Stockholm, Sweden*

| Cell type | HR (95%CI)* | P value | FDR |
| --- | --- | --- | --- |
| Leukocyte (10<sup>9</sup>/L) | 1.06(0.84-1.34) | 0.62 | 0.79 |
| Neutrophil (10<sup>9</sup>/L) | 1.07(0.85-1.34) | 0.58 | 0.79 |
| Lymphocyte (10<sup>9</sup>/L) | 1.04(0.82-1.31) | 0.76 | 0.79 |
| Monocyte (10<sup>9</sup>/L) | 0.97(0.76-1.23) | 0.79 | 0.79 |

\*Cox model was applied to derive the hazard ratios (HRs) with 95% confidence intervals (CIs) of risk of death, per standard deviation increase of the cell markers, with adjustment for age at diagnosis, sex, site of onset, diagnostic delay, ALSFRS-R score, time difference between the measure of ALSFRS-R score and diagnosis, BMI, and time difference between the measure of BMI and diagnosis.
FDR: false discovery rate.

**Supplementary Fig. 1.**
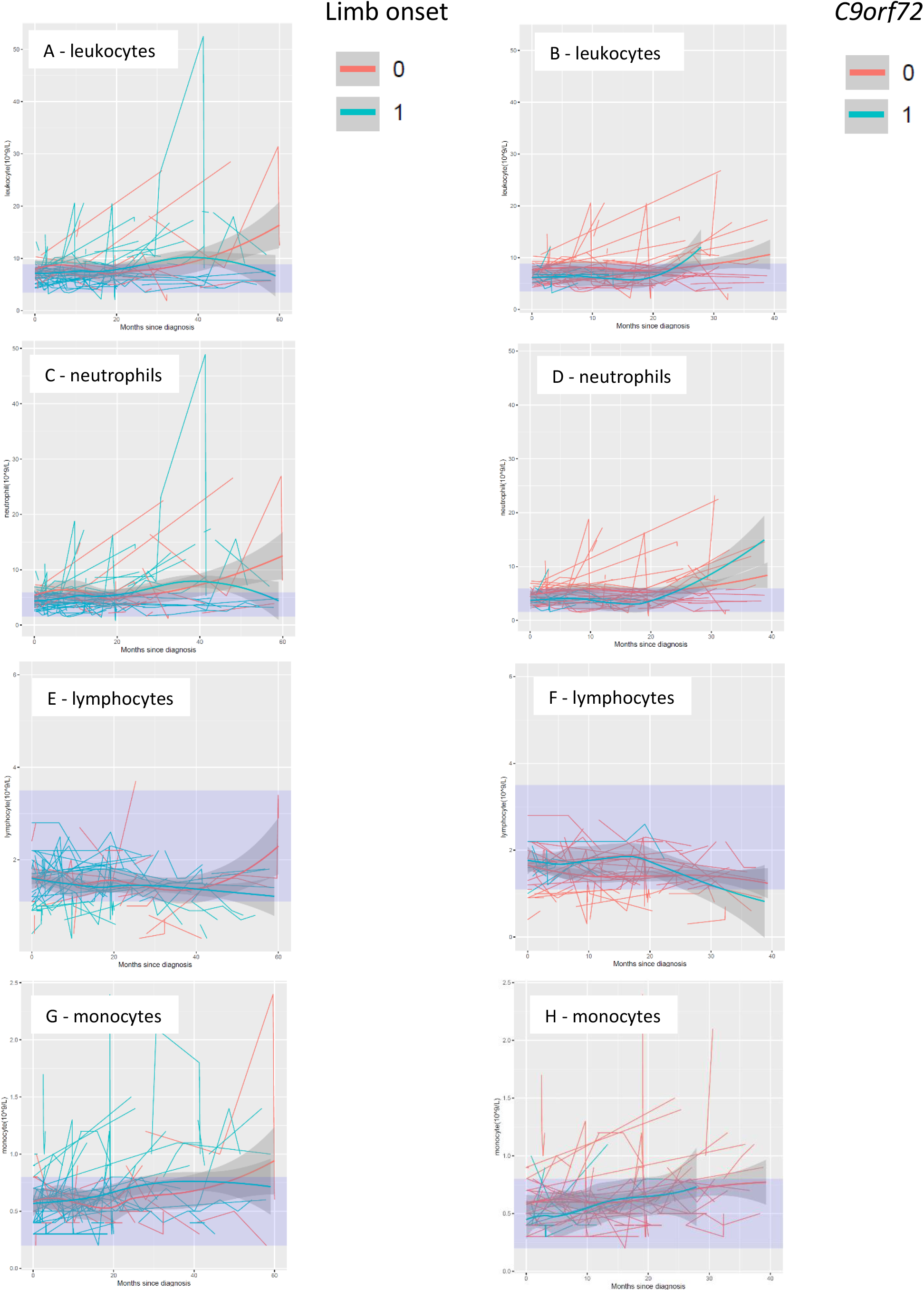
Temporal trend of leukocyte populations by site of onset and presence of *C9orf72* expansions.

**Supplementary Fig. 2.**
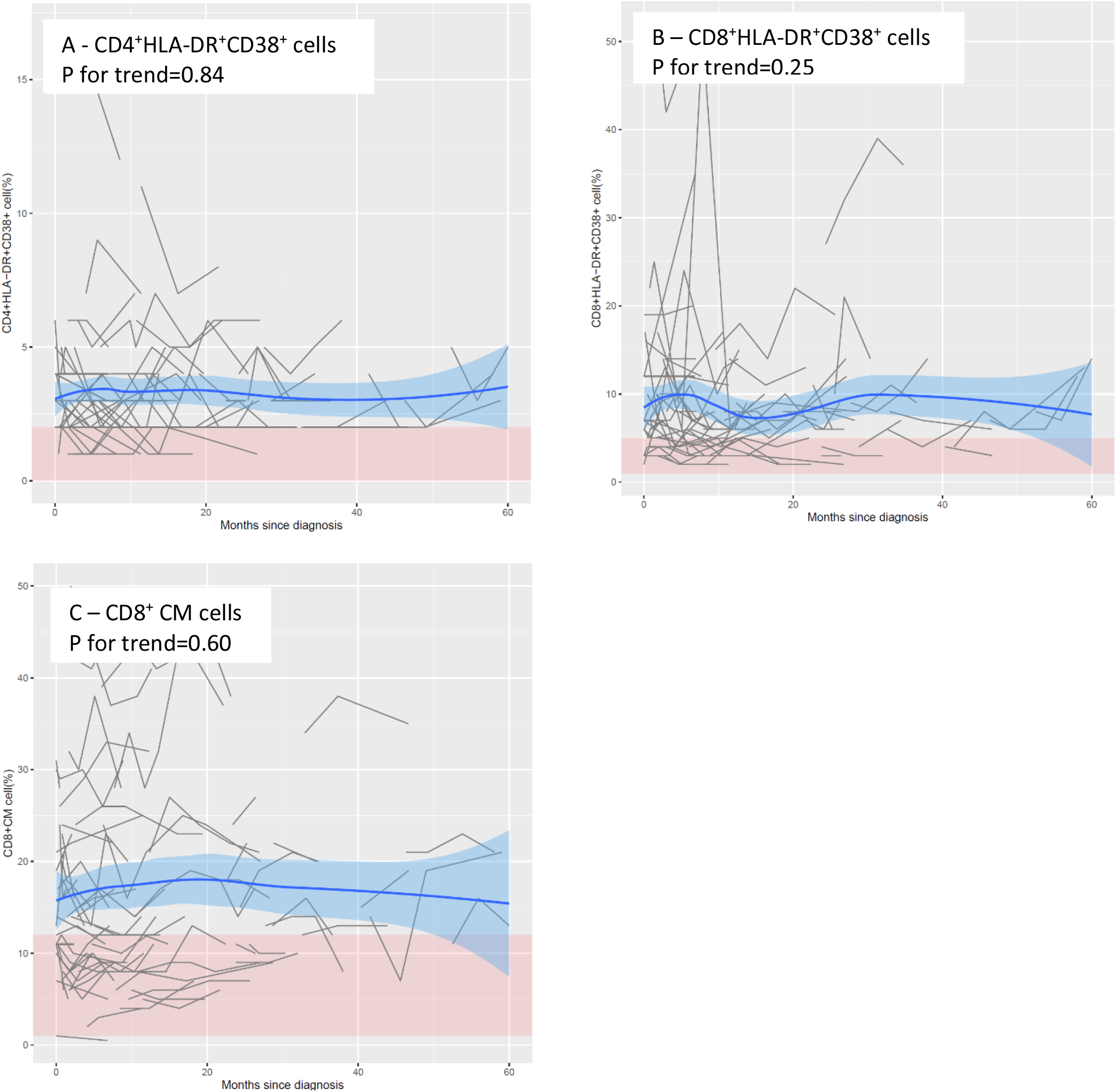
Lymphocyte populations that differed from normal range.

**Supplementary Fig. 3.**
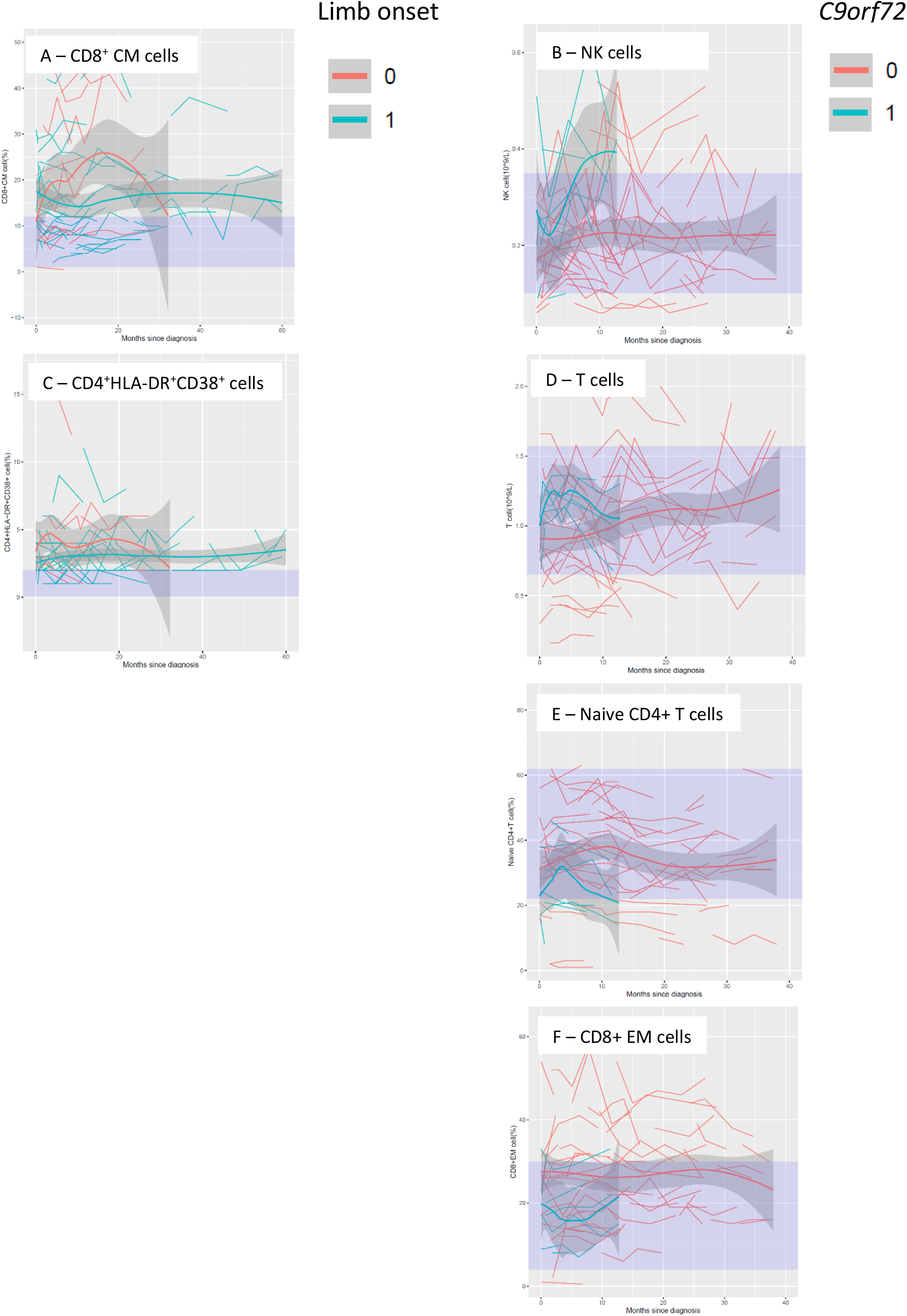
Temporal trend of lymphocyte populations that differed by site of onset and presence of *C9orf72* expansions.

**Supplementary Fig. 4.**
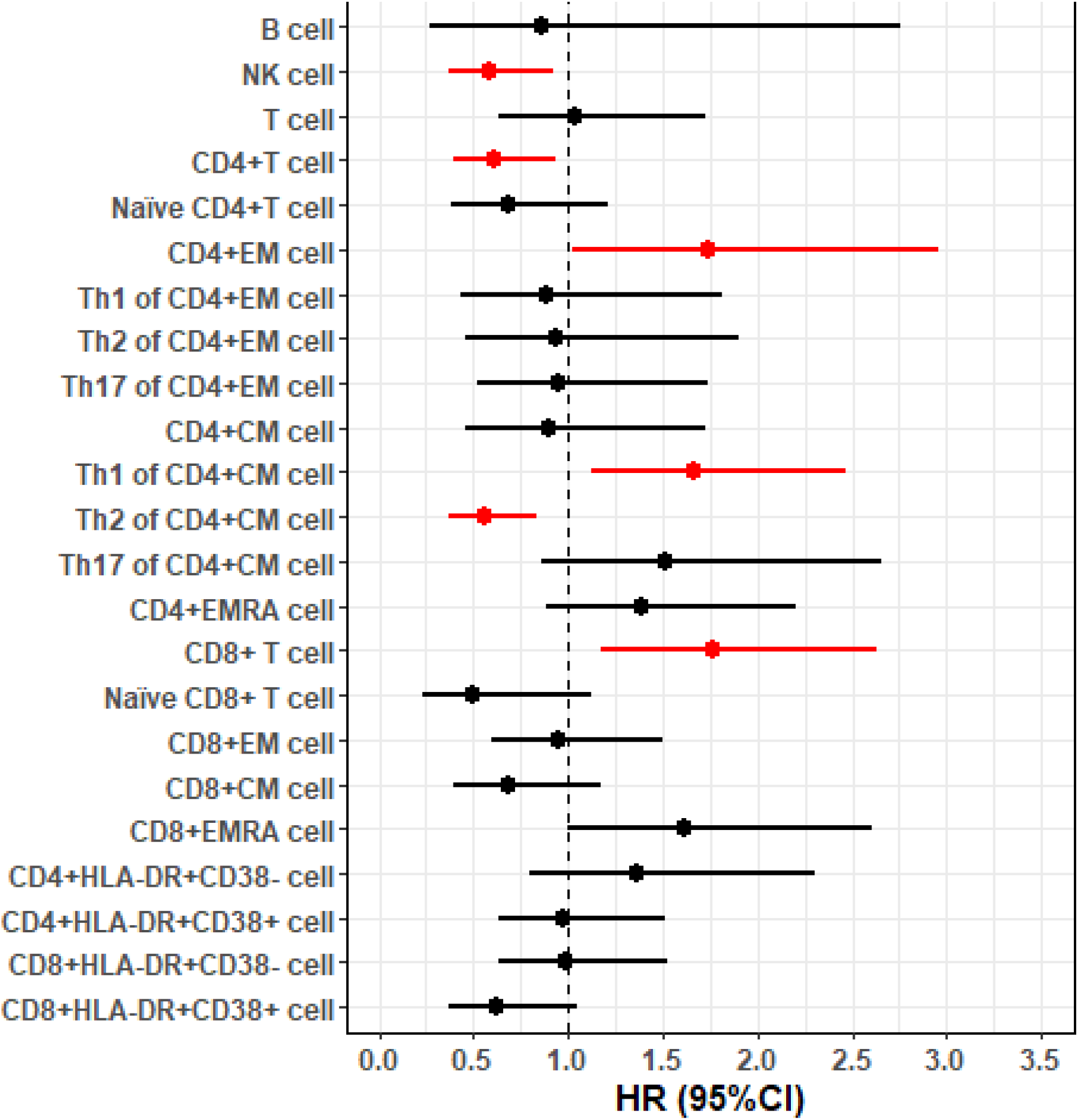
The associations of lymphocyte populations with risk of death after a diagnosis of amyotrophic lateral sclerosis (ALS), after excluding patients with *C9orf72* expansions.

